# Assessing fecal contamination from human and environmental sources using *Escherichia coli* as an indicator in rural eastern Ethiopian households - a study from the EXCAM project

**DOI:** 10.1101/2024.08.21.24312392

**Authors:** Loïc Deblais, Belisa Usmael Ahmedo, Amanda Ojeda, Bahar Mummed, Yuke Wang, Yitagele Terefe Mekonnen, Yenenesh Demisie Weldesenbet, Kedir A. Hassen, Mussie Brhane, Sarah McKune, Arie H. Havelaar, Song Liang, Gireesh Rajashekara

## Abstract

Enteric pathogens are a leading causes of diarrheal deaths in low- and middle-income countries. The Exposure Assessment of *Campylobacter* Infections in Rural Ethiopia (EXCAM) project, aims to identify potential sources of bacteria in the genus *Campylobacter* and, more generally, fecal contamination of infants during the first 1.5 years of life using *Escherichia coli* as indicator. A total of 1,310 samples (i.e., hand rinses from the infant, sibling and mother, drinking and bathing water, food and fomite provided to or touched by the infants, areola swabs, breast milk and soil) were collected from 76 households between May 2021 and June 2022. Samples were assigned to two groups by infant age: TP1 (time point 1), infants between 4 and 8 months of age, and TP2, infants between 11 and 15 months of age. Fluorometric and semi-selective colorimetric approaches were used to quantify *E. coli* in the field samples. Overall, *E. coli* was ubiquitous within selected households (56.8% across the study). *E. coli* was more frequently detected than average (>53%) with high concentration (>2-log CFU) in soil (g) and per pair of hand, while the opposite trend (<33%; <1.5-log CFU) was observed in food provided to the infants (g or ml), per areola, and breast milk (ml; P<0.01). *E. coli* was frequently detected in fomites touched by the infants, drinking and bathing water (>51%), but at low concentration (<1.5-log CFU). Correlation analysis between *E. coli* concentration in different sample types suggested that the mother’s hands might play a key role in the transmission of *E. coli* between the environment (i.e., soil, bathing water and fomites) and other family members (i.e., infant and sibling; P<0.04; r^2^>0.3). Using *E. coli* as surrogate, our study identified mother (hands and areola) as reservoirs likely to be involved in frequent transmission of fecal contaminants to infants within rural Ethiopian households.

## Introduction

Many infectious diseases can affect the human gastrointestinal tract, with enteric pathogens posing a significant health risk. Enteric infections often impair the gut’s intestinal integrity and function, leading to immediate adverse health consequences (e.g., diarrhea) and long-term chromic gastrointestinal issues (e.g., irritable bowel syndrome [IBS], inflammatory bowel disease [IBD], and chronic diarrhea) [1]. When combined with nutritional deficits during infancy, chronic exposure to enteric pathogens is linked to prolonged detrimental health conditions such as Environmental Enteric Dysfunction (EED) and stunting [2]. Conditions caused by EED are largely irreversible and lead to poor long-term health outcomes (e.g., diminished cognitive and physical development, weakened immune systems, and an increased risk of degenerative diseases later in life), especially when detected in young infant (<2-year-old) [3]. Enteric infections are also the primary causes of diarrhea, which is a leading cause of mortality and morbidity among children under five years old worldwide. These infections are particularly prevalent in resource-limited settings, due to limited access to clean water, inadequate sanitation, and poor personal hygiene practices. In 2017, diarrheal diseases were responsible for 1.6 million deaths globally, with the highest mortality rates primarily in Sub-Saharan Africa [4, 5]. Despite developmental efforts in these regions, access to clean water and proper sanitation management/infrastructure is precarious. Yet the evidence between enteropathogenic infections and related gastrointestinal disease is alarming, warranting global health interventions. This concern ultimately led the United Nations to include Clean Water and Sanitation as a Sustainable Development Goals (SDG-6) [6].

Climate change, food security, poor infrastructures, and inadequate water, sanitation, and hygiene (WASH) practices, contribute to a high disease burden in low and -middle-income countries (LMIC). Open defecation is a practice commonly performed by Ethiopian households and can lead to fecal-oral diseases such as diarrhea [7–9]. Global studies such as Etiology, Risk Factors, and Interactions of Enteric Infections and Malnutrition and the Consequences for Child Health and Development (MAL-ED) [10], WASH Benefits [11], and Sanitation Hygiene Infant Nutrition Efficacy (SHINE) [12] studies have highlighted the intricate interplay between these factors and their impact on health outcomes. These complex relationships present a significant challenge for low and -middle-income countries (LMIC) due to limited resources and inherent vulnerabilities to adapt to dramatic climatic shifts. Similar trends were observed in 2018 when our team conducted a cross-sectional study in rural Eastern Ethiopia as part of the CAGED project (*Campylobacter* Genomics and Environmental Enteric Dysfunction), during which WASH-oriented questionnaires revealed that only 23% of households used improved sanitation facilities shared between households and roughly 80% of households practiced open defecation [13]. In addition, most of the study population did not have access to clean water. Only 2% reported having access to a safely managed drinking water source on premises, available when needed, and not contaminated by fecal and chemical pollutants [13]. Diarrhea is often the most common symptom associated with a gastrointestinal infection, frequently characterized by a concentration of enteric pathogens disrupting the normal flora within the gut. Enteropathogenic microorganisms stem across different microbial groups, including bacteria (enteropathogenic or enterotoxigenic *E*. *coli*, *Shigella* spp., *Salmonella* spp., and *Vibrio cholerae*), parasites (such as *Cryptosporidium* spp. Giardia, and *Entamoeba histolytica*) and viruses (such as rotavirus, norovirus, astrovirus, and adenovirus). Given the numerous possible pathogens present at the point of fecal contamination, scientists rely on detecting fecal indicators such as bacterial coliforms for routine quality and safety monitoring of drinking water sources [14]. Testing for fecal indicators, such as *E. coli*, a member of the *Enterobacteriaceae* family within the Gamma proteobacterium class, has been crucial in identifying potential sources of contamination in water sources, particularly in resource-limited settings where enteric infections are highly prevalent [15, 16]. Furthermore, *E. coli* detection is a valuable tool because associated detection methods are relatively quick, easy, and inexpensive for routine quality and safety monitoring [17, 18].

In this study, we investigated the prevalence and concentration of *E. coli* in households from rural Eastern Ethiopia where newly born infants are raised, aiming to assess the potential level of fecal contamination surrounding the infant and identify potential fecal transmission pathways. This study is part of the Exposure Assessment of *Campylobacter* Infections in Rural Ethiopia (EXCAM) project, which aims to systematically evaluate infants’ exposure to *Campylobacter* and fecal contamination in rural districts of Haramaya, Eastern Ethiopia. *E. coli* was detected and quantified from a broad range of human (i.e., hand rinse, areola swabs, and breast milk) and environmental samples (i.e., drinking water, bathing water, food given to the infants, fomites touched by the infants and soil within the households) likely to be in close contact with infants.

## Materials and methods

### Study area and sample collection

The EXCAM study, nested within the CAGED project in the Eastern rural districts of Ethiopia [13, 19–21], recruited newborns from 79 households across 10 kebeles. The EXCAM study was based on similar methodology used in the SaniPath study [22]. Samples were collected during two distinct time windows: first, when infants were between 4 and 8 months of age (Time Point [TP] 1, between May 2021 and December 2021), and second, when they were between 11 and 15 months of age (TP 2, between December 2021 and June 2022). This approach allowed for the analysis of how behavior-mediated exposures may change with age during critical developmental stages. Additional details about the study area are previously described [13, 19–21]. Initially, 79 infants were included in TP1, decreasing to 76 in TP2 due to dropouts. Among the 76 infants who completed this study, 41 (54%) were male and 35 (46%) were female.

A total of 1,310 samples were collected across surveyed households between May 2021 and June 2022. During TP1, 586; samples were collected from infants under 8 months of age, while in TP2, 724 samples were collected from infants when they reached 11 to 15 months. Between 101 and 265 samples were collected by sample type during this study, with an average of 17 samples per household and 131 samples per kebele. Breast milk samples were mostly collected at TP1 because most mothers stopped giving breast milk when the infants were above 11 months of age. Soil samples collected inside the households were only collected at TP2 (n=57) because the infants motility increase after 11 months of age, and thus, allowing them to move around. Across the two periods, a total of 429 hand rinses, 117 breast milk samples, 149 areola swabs from the mother, 148 drinking water, 44 bathing water, 101 foods consumed by the infants, 265 fomites and 155 soil samples were collected. A total of 916 and 394 samples were collected during the dry season (between October and May) and rainy season (between June and September), respectively.

Three hand rinse samples (infant, sibling, and mother) per household per time point were collected by washing the hands of the surveyed individuals into a 1 L plastic bag prefilled with 200 ml of sterile distilled water. Hands were washed using normal hand-to-hand frictions for 30 seconds inside the bag. Areola swabs were collected using cotton swabs (Sterile Cotton Tipped Applicators, Puritan Medical Products; Guilford, ME) humified in sterile water. The humified swab was applied by the mother? directly to the skin, rotating it in a circular pattern on both areolas. The inoculated swab was transferred to a 15 ml tube prefilled with 4 ml Letheen broth (Difco BD; Becton, NY). Once the areola swab was collected, the areolas were cleaned with 70% ethanol to remove potential surface contaminants before collecting the breast milk. The breast milk samples were manually collected by the mother by applying pressure with her hand. The breast milk (at least 1 ml per mother per time point) was collected into a sterile 60 ml specimen collection cup (Starplex Scientific; Etobicoke, Canada). It should be noted that the mothers might not always have properly sanitized their hands before milk collection, potentially introducing contaminants into the milk. One liter of drinking and bathing water that was stored in a larger container in the household (e.g., bottles and jerrycans) were aseptically collected into four 532 ml Whirl-pak bags (Nasco WHIRL-PAK, Boston, MA) for each household at both time points. Foods (at least 10 g per household per time point) consumed by the surveyed infants was collected in a sterile 60 ml screw-capped container (Starplex Scientific; Etobicoke, Canada) using the utensil (e.g., spoon, fingers) commonly used to manipulate the food and feed the infant. Surfaces (aka. fomites) the most frequently touched by the surveyed infants were swabbed using a Speci-sponge (Nasco WHIRL-PAK; Boston, MA) pre-humified with 10 ml of sterile water. The selection of three mostly touched fomites by the infant of the designated households was based on the total area touched by the infant (e.g., floor, carpet, or phone). Behavioral data related to these fomites for the surveyed infants are detailed in a companion paper (Wang et al., 2024, in preparation). The designated area was swabbed by moving the humected Speci-sponge back and forth at least five times on both sides. The inoculated sponge was transferred back into its original collection bag.

Three soil samples per household were collected inside (one sample where the floor is covered by a carpet or plastic sheet, and one sample where the floor is made of dust and dirt) and outside (one sample collected in front of the house) using boot socks humified with 207 ml of 10% skim milk. Each soil sample was collected by taking 15 steps within the designated area (preferably a 3 x 5 m area where children have access making sure to cover most of the area). Then the boot socks were removed using aseptic techniques and transferred into the original Whirl-pak bag humified with 10% skim milk. All samples described above were stored and transported on ice to the lab for processing. Phosphate buffered saline solution or equivalent could not be used to collect the field samples (i.e., hand rinse, areola swab and fomites) instead of sterile water because the surveyed households did not believe the buffer was healthy to be touched, especially for the infant.

### Sample Processing

Hand rinse, drinking, and bathing water samples were designated as large, diluted samples, and thus, concentrated using 500 mL Disposable 0.22 µm Nylon Vacuum Filter (Corning; Corning, NY). The membrane was aseptically removed and cut into 5 mm^2^ pieces using sterile blade and forceps. The cut membrane was transferred into a 5 ml tube pre-filled with 4 ml of 1X peptone water (pH 7) and twenty 3-mm diameter glass beads. The sample was manually homogenized for at least 1 min to release cells from the nylon membrane. The supernatant was transferred into another 5 ml tube and centrifuged for 10 min at 4,500 rpm. The pellets were concentrated into 1 ml of 1X peptone water (pH 7).

Soil samples (boot socks), fomites, and areola swabs were identified as dry samples, and thus, resuspended into 25, 10, and 3 ml of 1X peptone water (pH 7), respectively. Resuspended samples were homogenized for 1 min by hand for bags or using a vortex for tubes. The liquid obtained for the homogenization was transferred into 50 ml tubes and centrifuged for 10 min at 4,500 rpm. The pellets were carefully concentrated into 1 ml of 1X peptone water (pH 7). Liquid food samples (e.g., cow milk and mango jus) and breast milk samples were used as is. One gram of solid food samples (e.g., biscuit, rice, spaghetti, macaroni and injera) was manually homogenized for 30 sec into a 100 ml Whirl-Pak bag (Seattle, WA) filled with 9 ml of 1X peptone water (pH 7). The homogenize samples were centrifuged for 10 min at 4,500 rpm and the pellet were resuspended into 1X peptone water (pH 7; 5 ml final volume).

### Detection and quantification of *Escherichia coli* in clear and translucent field samples using EC-MUG fluorometric approach

The *E. coli* concentration in clear (e.g., drinking and bathing water) and translucent (e.g., hand rinse and fomites) samples was estimated using the modified MPN 96-well plate assay combined with EC-MUG fluorometric approach. Based on the total volume available per sample’s type, a total of 15.84 ml of sample (n=88 reactions; 180 µl of the designated sample per well = one reaction) was tested from hand rinse, drinking and bathing water samples (also called “large liquid samples”), while 0.9 ml of concentrated samples (n=5 reactions; 180 µl of the designated sample per well = one reaction) were tested from fomite, areola swab and food samples (also called “small liquid samples”). For both “large and small liquid samples”, 1.62 ml of EC-MUG broth was added into each well of a 96-deep well plate (VWR; Radnor, PA). In addition, four wells were inoculated with 180 µl of sterile 1X peptone water (pH 7; negative controls) or four wells were inoculated with 180 µl of an overnight *E. coli* culture known to produce fluorescence (positive controls). The inoculated plate was sealed using adhesive plate seal (Thermo Scientific; Waltham, MA) and incubated at 37°C for 48 hrs. After incubation, 200 µl of incubated product was transferred into a transparent 96-well plate (VWR; Radnor, PA) and expose to UV light to enumerate number of fluorescence wells (indicating the presence of *E. coli*). In parallel, the field samples were serially 10-fold diluted up to 8 times in EC-MUG broth in case if the *E. coli* concentration was too high to be quantified using the first approach. For the analysis of the data, we hypothesized that one positive well harbors at least one viable *E. coli* CFU.

### Detection and quantification of *Escherichia coli* population in turbid/opaque field samples using Chromocult colorimetric approach

Opaque samples (Breast milk, soil and opaque food samples) were 10-fold serial diluted in 1X peptone water (pH 7). Two hundred microliters of undiluted and diluted samples were plated on Chromocult® Coliform agar (Sigma Aldrich Millipore; Saint-Louis, MO). The plates were incubated at 37°C for up to 48 hrs. Colonies with purple color (characteristic of *E. coli*) were recorded.

### Statistical analysis

*E. coli* concentration data were log-transformed. Statistical analyses were performed using JMP PRO 16 software (SAS Institute; Cary, NC, USA) and R v4.2.1 (https://cran.r-project.org/). The normality of the *E. coli* concentration data was assessed using Goodness of fit combined with Shapiro test. Chi^2^ and Wilcoxson tests were used to identify differences in *E. coli* prevalence and concentration, respectively based on the recorded metadata (e.g., kebeles, infant sex, samples types, and collection dates). A multivariate analysis (Pearson correlation and linear regression reported for each pairwise comparison axis) was used to identify correlation between the concentration of *E. coli* and sample types for the designated time points. A P-value of 0.05 was used at the cut-off for the above statistical tests.

## Results

### Prevalence of *E. coli* in the Ethiopian households

Overall, *E. coli* was detected in 58% (745/1,310) of all the samples, and no significant differences in *E. coli* prevalence were detected between TP1 (58% [n= 341/586; CI95%= 54-62%]) and TP2 (56% [n= 404/724; CI95%= 52-59%]; P>0.05; **Table 1**). No difference in *E. coli* prevalence was detected across the 10 kebeles surveyed in this study (56% ±9.2% and 55% ±6.2% at TP1 and TP2, respectively; **Table 1**).

**Table 1.** Prevalence of *Escherichia coli* in the ten kebeles surveyed in rural eastern Ethiopia.

| **Kebele ID** | **Time point 1** |  |  |  | **Time point 2** |  |  |  |
| --- | --- | --- | --- | --- | --- | --- | --- | --- |
|  | **Sample size** | **Mean** | **Lower 95%** | **Upper 95%** | **Sample size** | **Mean** | **Lower 95%** | **Upper 95%** |
| 1 | 73 | 56.2 | 44.5 | 67.8 | 96 | 52.1 | 41.9 | 62.2 |
| 2 | 83 | 59 | 48.2 | 69.8 | 97 | 56.7 | 46.7 | 66.7 |
| 3 | 51 | 62.7 | 49 | 76.7 | 66 | 56.1 | 43.8 | 68.3 |
| 4 | 49 | 57.1 | 42.8 | 71.5 | 57 | 54.4 | 41.1 | 67.7 |
| 5 | 56 | 58.9 | 45.6 | 72.2 | 74 | 52.7 | 41.1 | 64.3 |
| 6 | 81 | 61.7 | 50.9 | 72.5 | 93 | 53.8 | 43.4 | 64.1 |
| 7 | 78 | 55.1 | 43.8 | 66.4 | 92 | 60.9 | 50.7 | 71 |
| 8 | 13 | 30.7 | 17.3 | 60 | 20 | 40 | 16.5 | 63.5 |
| 9 | 60 | 61.2 | 49 | 74.3 | 78 | 62.8 | 51.8 | 73.8 |
| 10 | 42 | 57.1 | 41.5 | 72.7 | 51 | 56.9 | 42.8 | 70.9 |
| Total | 586 | 58.2 | 54.2 | 62.2 | 724 | 55.8 | 52.2 | 59.4 |

Value in percentage.

*E. coli* prevalence was divided into two distinct groups based on the sample types studied (e.g., prevalence below 33% versus higher than 53%; P<0.001; **Figure 1A**). Specifically, *E. coli* was more frequently detected in bathing water (91% [CI95%= 79-100%] at TP1 and 81% [CI95%= 63-99%] at TP2), drinking water (76% [CI95%= 66-86%] at TP1 and 51.3% [CI95%= 40-63%] at TP2), fomites touched by the infants (62% [CI95%= 54-71%] at TP1 and 56% [CI95%= 48-65%] at TP2), mother hand rinse (80% [CI95%= 72-89%] at TP1 and 84% [CI95%= 76-93%] at TP2, respectively), sibling hand rinse (88% [CI95%= 81-97%] at TP1 and 85% [CI95%= 76-94%] at TP2), infant hand rinse samples (52% [CI95%= 41-64%] at TP1 and 78% [CI95%= 68-87%] at TP2), and soil samples collected inside the households (100% at TP2) compared to food eaten by the infants (33% [CI95%= 16-50%] at TP1 and 25% [CI95%= 14-36%] at TP2), areola swabs (16% [CI95%= 7-24%] at TP1 and 19% [CI95%= 10-28%] at TP2) and breast milk samples (21% [CI95%= 9-33%] at TP1 and 3% [CI95%= 0-7%] at TP2).

**Figure 1.**
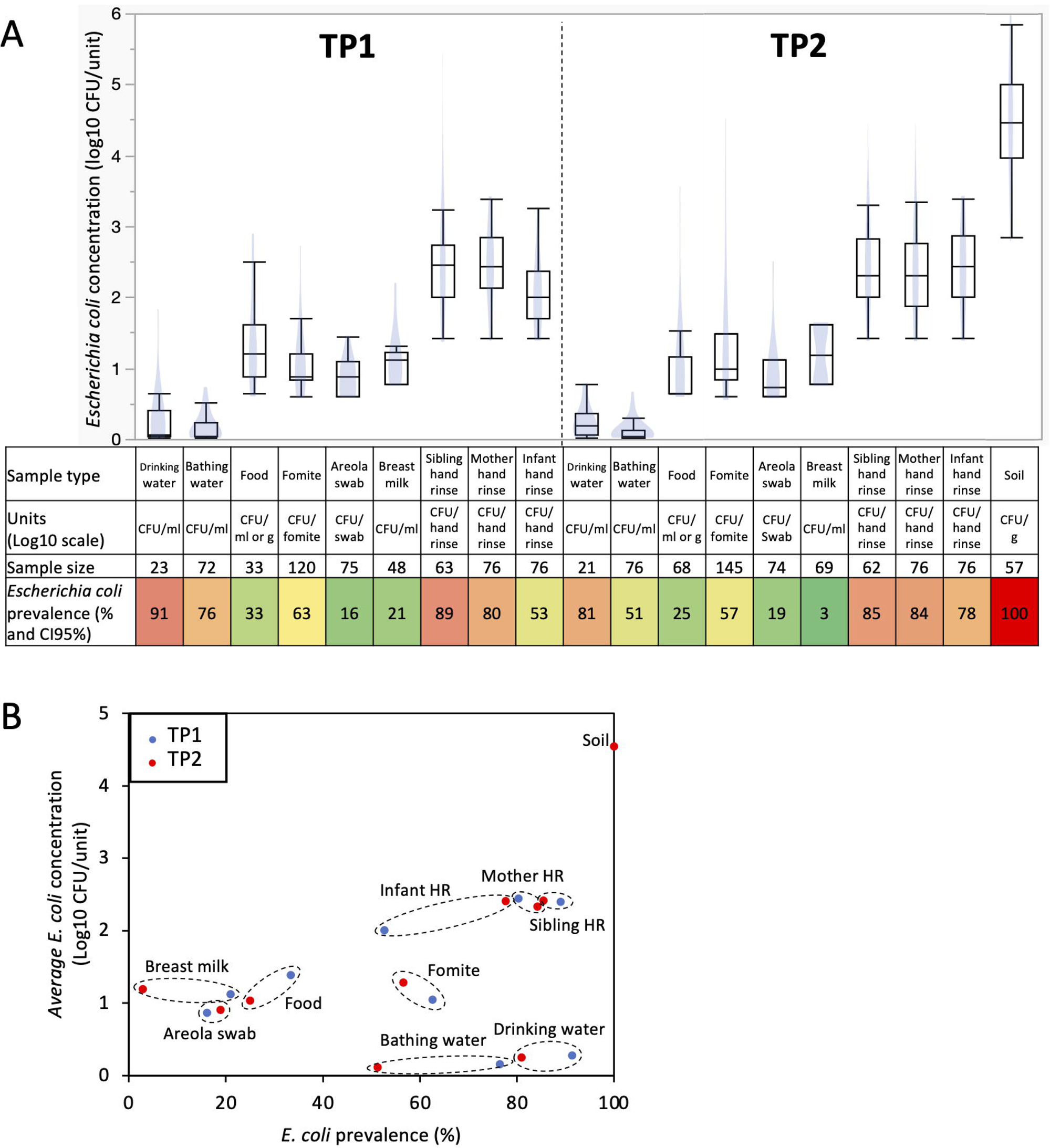
Prevalence and concentration of *Escherichia coli* in samples collected within 76 Ethiopian households between May 2021 and June 2022. A) Boxplots of *E. coli* concentration and table showing *E. coli* prevalence. Concentration data were determined only based on the samples positive for *E. coli*. Blue shadows within the boxplots represents the density ellipse (graphical representation that can show data extent, center of mass, least squares fit line and outliers) obtained based on the *E. coli* concentration data for the designated sample type and time point. HR: hand rinse. The color of the cells in the table are proportional to the *E. coli* prevalence data (e.g., red means high prevalence and green means low prevalence). B) Comparison between *E. coli* prevalence and concentration. Samples from TP1 (red dots; n=586; between May 2021 and December 2021) were collected when the infants were under 8 months of age, while the samples from TP2 (blue dots; n=724; between December 2021 and June 2022) were collected when the infants between 11 and 15 months of age. Dotted circles cluster data (*E. coli* prevalence and concentration) obtained for similar sample type at TP1 and TP2.

*E. coli* prevalence decreased across most sample types at TP2 compared to TP1 (c.a., 12% ±8 decrease; especially in bathing and drinking water, and breast milk; P<0.05; **Figure 1A**), while the opposite trend was observed with the infant hand rinse (c.a., 25% increase; P<0.002). Furthermore, variations in *E. coli* prevalence were also observed based on the date of sample collection and the age of the infants when the samples were collected. The prevalence of *E. coli* in the infant hand rinse significantly increased based on the infants’ age at sampling (r^2^=0.2; P=0.006). These results were not affected by the sex of the infant. Overall, 86% and 88% of the hand rinse samples collected from male and female infants were positive for *E. coli*, respectively. Other sample types also displayed equivalent *E. coli* prevalence between sex (P>0.05). On the other hand, the opposite trend was observed with drinking water and breast milk samples. The prevalence of *E. coli* in the drinking water and breast milk significantly decreased over time (r^2^=−0.2 and −0.3, respectively; P<0.004) and significantly decreased over time based on the age of the infant when the samples were collected (r^2^=−0.2 and −0.3, respectively; P<0.006).

No differences in *E. coli* prevalence were detected between the food types provided to the infants (e.g., biscuit soaked in water, cow milk, injera, Fanta soda, macaroni, sliced mango, and boiled rice and corn) and the fomites touched by the infants (e.g., bottle used to feed the infant, mobile phone, plastic bag, and teacups). However, it is worth nothing that the sample size of the mentioned food items was small, which affected the power of the statistical analysis. *E. coli* was the most frequently detected in food items consumed by the infants (i.e., n= 3/6 cooked rice, 2/5 macaroni, 4/9 cow milk and 10/45 injera) and fomites touched by the infants (i.e., n= 5/7 mobile phone and 6/15 tea cup).

### Concentration of *E. coli* in the Ethiopian households

The concentration of *E. coli* discussed in this section is based only on the *E. coli* positive samples. Overall, *E. coli* was detected in 745 samples processed in this study (n= 341 and 404 positive samples at TP1 and TP2, respectively). No difference in *E. coli* concentration was detected between kebeles and households for the sample types studied. On the other hand, the concentration of *E. coli* was separated into four distinct groups across the sample type (**Figure 1A**). Drinking and bathing water harbored low level of *E. coli* (<0.3-log CFU/ml), followed by foods eaten by the infants, fomites touched by the infants, areola swabs and breast milk with a concentration of *E. coli* between 0.9- and 1.2-log CFU/ml, g or sample (i.e., areola swabs and fomites touched by the infants; **Figure 1A**). Interestingly, the hand rinse samples collected from the mother, sibling and infant harbored an average concentration in *E. coli* between 2.2- and 2.4-log CFU/pair of hands (**Figure 1A**). Finally, *E. coli* was the most abundant in the soil samples (4.5-Log CFU/g ±0.1). As observed with the prevalence data, the *E. coli* population from the hand rinse collected from the infants was positively correlated with the date of collection (r^2^=0.3; P=0.002) and the age of the infant when the samples were collected (r^2^=0.3; P=0.001). Therefore, *E. coli* was significantly higher in the hand rinse collected from the infant at TP2 (2.4-log CFU/fomite ±0.1; n=59 samples positive for *E. coli*) compared to the ones collected at TP1 (2-log CFU/fomite ±0.1; n=40 samples positive for *E. coli*; P=0.001; **Figure 1A**). These results were not affected by the sex of the infant. Overall, 2.4-log *E. coli* CFU/hand rinse were detected from both male and female infants hand rinse samples, respectively. Other sample types also displayed equivalent *E. coli* prevalence between sex (P>0.05). *E. coli* concentration was also significantly higher in fomites touched by the infants collected at TP2 (1.3-log CFU/fomite ±0.1; n=82 samples positive for *E. coli*) compared to the ones collected at TP1 (1.1-log CFU/fomite ±0.1; n=75 samples positive for *E. coli*; P=0.03; **Figure 1A**).

The concentration of *E. coli* was not always associated with the prevalence in the designated sample type (**Figure 1B**). High prevalence (>53%) and concentration of *E. coli* (>2-log CFU/sample) were detected in soil (gram) collected inside the households and the pair of hands. Low prevalence (<33%) and concentration of *E. coli* (<1.5-log CFU/sample) were detected in the foods (g or ml) eaten by the infants, areola and breast milk (ml; **Figure 1B**). On the other hand, *E. coli* was frequently detected in fomites touched by the infants, drinking and bathing water (ml; >51%), but at low concentration (<1.5-log CFU/sample; **Figure 1B**).

### Correlations between *E. coli* concentration in reservoirs in the Ethiopian households

Distinct correlations in *E. coli* concentration were observed between sample types at the two time points, TP1 (**Figure 2A**) and TP2 (**Figure 2B**). At TP1, when the infants were under 8 months of age, most of the correlations were centralized around the hand rinse samples collected from the household members (i.e., mother, sibling and infant; green area in **Figure 2A**). Specifically, the increase concentration of *E. coli* detected in the hand rinse from the mother was positively correlated with the concentration of *E. coli* from the infant hand rinse (r^2^=0.5; P=0.0001) and the sibling hand rinse (r^2^=0.4; P=0.002; green area in **Figure 2A**). To some extent the concentration of *E. coli* between the infant hand rinse and the sibling hand rinse were positively correlated (r^2^=0.3; P=0.02; green area in **Figure 2A**). Another set of the correlations were centralized around the interaction between the mothers and the infants (i.e., hands and areola swabs; pink area in **Figure 2A**). Both, the concentration of *E. coli* detected in the mothers’ and the infants’ hand rinses were positively correlated with concentration of *E. coli* in the areola swabs (r^2^=0.3 and 0.4, respectively; P<0.009), which was itself to positively correlated with the concentration of *E. coli* in the breast milk (r^2^=0.3; P=0.05; pink area in **Figure 2A**). Both the concentration of *E. coli* in the areola swabs and infant hand rinse were positively correlated with the fomites touched by the infants (r^2^=0.2 for both sample types; P=0.03; pink area in **Figure 2A**).

**Figure 2.**
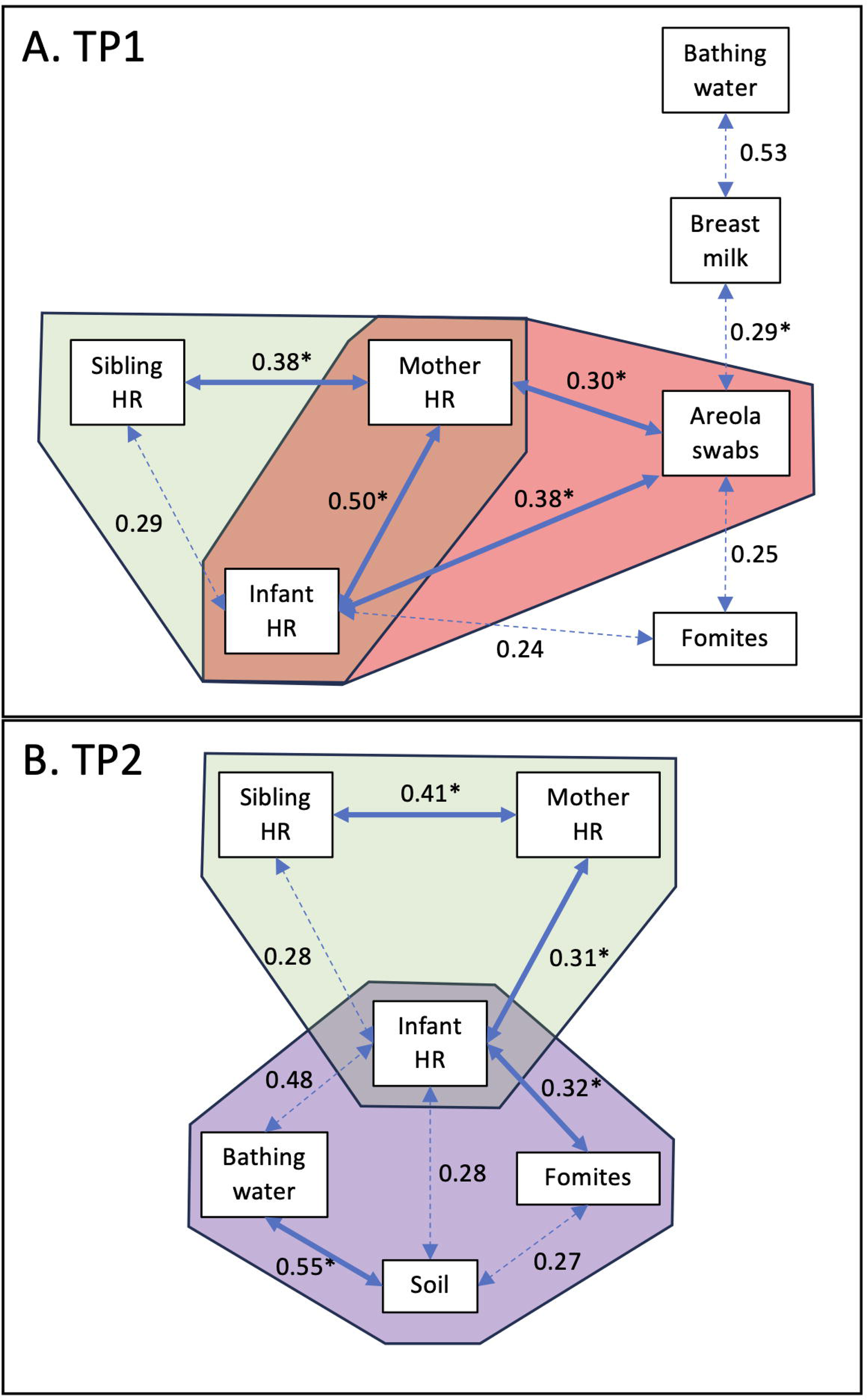
Correlation of *E. coli* concentrations between sample types. A) Correlation detected at TP1 when the infants were under 8 months of age. B) Correlation detected at TP2 when the infants were between 11 and 14 months of age. Values indicate the r^2^ values. The full arrows combined with the symbol “*” indicates that the p-value was lower than 0.01. The dotted arrows indicates that the p-value was lower than 0.05. Only significant data (P<0.05) were displayed in the network maps.

At TP2, when the infants were between 11 and 15 months of age, most of the correlations were centralized around the infants’ hand rinse samples (**Figure 2B**). As observed in TP1, the concentration of *E. coli* in the infants’ hand rinse was also positively correlated with the concentration of *E. coli* from the mothers’ hand rinse (r^2^=0.3; P=0.005), and to some extent to the siblings’ hand rinse (r^2^=0.3; P=0.03; green area in **Figure 2B**). The concentration of *E. coli* in the mother hand rinse was still positively correlated with the one in the sibling hand rinse (r^2^=0.4; P=0.008; green area in **Figure 2B**). The concentration of *E. coli* in the infants’ hand rinse was positively correlated with the concentration of *E. coli* from the fomites touched by the infants (r^2^=0.3; P=0.004; purple area in **Figure 2B**), and to some extent with concentration of *E. coli* in the soil collected within the households and the bathing water (r^2^=0.3 and 0.5, respectively; P=0.03; purple area in **Figure 2B**). The *E. coli* concentration in the soil samples collected inside the households was positively correlated with the bathing water (r^2^=0.5; P=0.01; purple area in **Figure 2B**) and to some extent with fomites touched by the infants (r^2^=0.3; P=0.04; purple area in **Figure 2B**).

## Discussion

The transmission of fecal contaminants within rural households poses a significant health concern [23, 24]. Poor sanitation and inadequate hygiene practices can increase exposure to fecal pathogens, like diarrheagenic *E. coli, Salmonella,* and *Campylobacter*, leading to diarrheal diseases and other health complications, especially among vulnerable infants [25]. This study analyzed commensal *E. coli* as an indicator of fecal contamination in 1,310 samples collected across two time points in the households of infants in rural Ethiopia. Seventy-nine households participated in the study of which 76 completed follow-up. Overall, *E. coli* was detected in 56.8% of samples collected between May 2021 and June 2022. Similar observations were reported from studies conducted in at least 27 countries in Africa (*E. coli* prevalence between 47% - 83%), involving both environmental and household samples [26–28]. *E. coli* was detected in all sample types. Hand rinse samples collected from the household members (i.e., mother, sibling, and infant) were most frequently contaminated with *E. coli* at high concentrations (>53% prevalence and >2.25-log CFU per set of hands). These findings are in alignment with previous studies reporting that hands are a recurrent carrier of fecal contaminant due to improper sanitation after defecation or contact with a contaminated environment [29–31]. *E. coli* was also frequently detected, albeit in lower concentration on fomites touched by the infants, and in their drinking and bathing water*. E. coli* concentration obtained in this study are equivalent to data reported from LMIC (e.g., Bangladesh, Uganda and Cambodia) [32]. On the other hand, breast milk, areola swabs and food given to the infants were less likely to have *E. coli.* Nevertheless, it is breast feeding is an action frequently conducted in order to feed infants. Statistic models incorporating the *E. coli* data with the observation data recorded during this study will be developed to determine whether reservoirs less likely to harbored *E. coli* or at low concentration still have a predominant risk for the transmission of E. coli and other fecal contaminants to the infants.

The presence of *E. coli* within specific reservoirs and its potential movement between sample types significantly varied with age (when the infants were below 8 months of age versus when the infants were between 11 and 15 months of age). The presence of *E. coli* on the infants’ hands was associated with their age (P<0.01). *E. coli* was less frequently detected when the infants were under 8 months of age (53%) compared to the hand rinse samples collected when the infants were between 11 and 15 months of age (78%). This increase in *E. coli* prevalence may be linked to the infants’ increased autonomy and their greater interaction with their environment. In Ethiopia, it is customary for mothers and infants to stay indoors together for extended periods post-birth, which may facilitate transfer of *E. coli* as the mothers’ hand contamination was most correlated with *E. coli* levels in the infant hands during the first period of this study (when the infants were under 8 months of age). Our data highlight the daily routine that the mothers are engaging with the infants could lead to repeated exposures of the infant with fecal contaminants, especially during breast feeding and handling the infant, as reported by previous studies [33, 34]. We previously showed that the enteric pathogen *Campylobacter* was highly prevalent in mothers’ stools (83.1%) collected from the same households [21]. It was also found that the *Campylobacter* concentration in the mothers’ stools correlated with the *Campylobacter* concentration in the infants’ stools [21], which supports the transfer of fecal contaminants between mother and infant.

Interestingly, only minor differences in *E. coli* concentration were detected over time, suggesting that seasonal variations did not significantly impact *E. coli* presence in the studied households. On the other hand, there were marked changes in potential *E. coli* transfer within the households between the first period (TP1) and the second period (TP2) of this study. As mentioned above, initially interactions observed in TP1 predominantly involved mothers (i.e., hand rinse and areola swabs) and the infants. During the second period, the infant became central to these interactions, not only maintaining strong interactions with their mothers’ hands but also increasingly engaging with other environmental reservoirs such as fomites, bathing water and soil within the households. This shift could be explained by the infants’ developmental progression and contact with various surrounding reservoirs. Similar observations were reported in our previous study conducted in the same region in eastern Ethiopia, where the prevalence and concentration of *Campylobacter* in the infants were closely correlated with the concentration of *Campylobacter* in the soil collected inside the households and presence of specific livestock (e.g., goat and sheep) [21]. It should be noted that when the infants reach 11 months of age, breast feeding becomes less frequent and with increasing exposure to other foods (e.g., injera, rice and dry biscuit), which are less likely to have *E. coli*. However, given all the households selected in this study owned livestock (i.e., chicken, cow, goat and/or sheep), the infants were more likely to encounter particles rich in fecal contaminants [35].

As previously described in our recent published studies [20, 21], this study faced several limitations during its execution and sample collection, which significantly influenced the methodology used to detect and quantify *E. coli*. A limited amount of drinking water (i.e., 1 L per household per time point) could be collected from the selected households due to the constraint by limited resources available and cultural norms. Further, hesitation from some mothers to provide samples was encountered due to stigma, rumors, and cultural norms. Due to the sensitive nature of the operation, areola swabs and breast milk collections were performed privately by the mother. Thereby, the quality and number of samples collected was likely to fluctuate between households. The goal of this study was to develop an easy, low-cost, and high-throughput tests that could be applied on clear/translucent samples (e.g., drinking and bathing water, hand rinse, fomites touched by the infants and areola swabs) or opaque samples (e.g., food provided to the infants, milk and soil collected from the households). The prevalence and concentration of *E. coli* reported in this study were determined based on standardized selective colorimetric Chromocult plating [36, 37] and fluorometric EC-MUG approaches [38, 39]. Thereby, the results indicated the total *E. coli* population but do not differentiate between commensal and pathogenic *E. coli*. Further, we acknowledge that the prevalence of *E. coli* in the drinking water might have been underestimated due to the limited sample volume processed in this study. Given the mothers’ hands harbored high level of *E. coli*, our study might have overestimated the prevalence of *E. coli* in the areola swabs and breast milk samples due to potential cross-contamination with the hands during the sample collection. The *E. coli* data will be compared to the behavioral data collected in the households surveyed in this study to determine whether these sample types are reservoirs associated with the frequent transmission of *E. coli* to the infants (Wang et al., in preparation).

In conclusion, our study revealed a high prevalence and concentration of *E. coli* in the different reservoirs surrounding infants, indicating significant fecal contamination and potential for transmission of associated enteric pathogens (e.g., *Campylobacter* spp., pathogenic *E. coli*, rotavirus, and *Salmonella enterica*) within rural Ethiopian households. Future studies will assess whether the prevalence and concentration of *E. coli* could be used as bioindicator for other enteric pathogens (i.e., *Campylobacter*) in these low-income resource settings. Furthermore, the interconnections detected between *E. coli* reservoirs highlighted different routes that could lead to transmission of fecal contaminants within the households, and thus, could potentially lead to early infection of the infants with enteric pathogens.

## Data Availability

Availability of data and materials. All data and materials used in this study are available upon reasonable request. The datasets generated and/or analyzed during the current study are not publicly available due to ethical restrictions, but can be made accessible through the corresponding author on reasonable request. Any additional information regarding the materials and methods is also available from the corresponding author upon request.

## Declaration

### Ethics approval and consent to participant

Ethical approval was obtained from the University of Florida Internal Review Board (IRB201903141); the Haramaya University Institutional Health Research Ethics Committee (COHMS/1010/3796/20) and the Ethiopia National Research Ethics Review Committee (SM/14.1/1059/20). Written informed consent is obtained from all participating households (husband and wife) using a form in the local language (Afaan Oromo). Research findings will be disseminated to community stakeholders, including participants, through the existing CAB. Findings will be disseminated to scientific, academic, policy and development stakeholders through conferences and peer-reviewed journals and through the Feed the Future Innovation Lab for Livestock Systems. The Bill and Melinda Gates Foundation, the funder of this trial, requires an open access data policy. Therefore, all manuscripts from this funded work will be open access with the data underlying the published research results available in a public repository. The website https://www.gatesfoundation.org/how-we-work/generalinformation/open-access-policy provides more information on this policy.

A Community Advisory Board including a representative of the community, religious leaders (imam), woreda and kebele administration, woreda women and children affairs, woreda bureau of health and agriculture, kebele health, and agricultural extension workers was established to guide the research team for better understanding of local context and entry to the community and is regularly engaged in the research [20]. Only the project manager at Haramaya University and the data manager at the University of Florida have access to personally identifiable information in the REDCap database. Any data shared among researchers within the project was identified and blinded. Materials and Data Transfer Agreements assure confidentiality of data when exchanged with international partners and others.

### Consent for publication

The authors confirmed that all relevant parties have reviewed the final version of the manuscript and have provided their explicit consent for its publication. All contributors and individuals involved in the research have given their informed consent for the publication of their data in this article.

### Availability of data and materials

All data and materials used in this study are available upon reasonable request. The datasets generated and/or analyzed during the current study are not publicly available due to ethical restrictions, but can be made accessible through the corresponding author on reasonable request. Any additional information regarding the materials and methods is also available from the corresponding author upon request.

### Competing interests

The authors declare that they have no competing interests.

### Funding

This project is funded by the United States Agency for International Development Bureau for Food Security under Agreement #AID-OAA-L-15-00003 as part of Feed the Future Innovation Lab for Livestock Systems, and by the Bill & Melinda Gates Foundation OPP#1175487. Under the grant conditions of the Foundation, a Creative Commons Attribution 4.0 Generic License has already been assigned to the Author Accepted Manuscript version that might arise from this submission. Any opinions, findings, conclusions or recommendations expressed here are those of the authors alone. Research reported in this publication was supported by the University of Florida Clinical and Translational Science Institute, which was supported in part by the NIH National Center for Advancing Translational Sciences under award number UL1TR001427.

### Author’s contributions

LD, BUA, AO and GR wrote the main manuscript text. LD, BUA, AO, were involved in the analysis of the data. LD, BUA, AO, YW, were involved in the interpretation of the data. LD, BUA, AO, BM, YTM, YW, YDW, KAH, and MB were involved in the acquisition of the data. SM, AHH, SL, LD and GR were involved in the conception and design of the work. All authors reviewed the manuscript

## Acknowledgements

We thank the local community advisory board for facilitating this research in different selected kebeles. The EXCAM project is funded by the Bill & Melinda Gates Foundation to address food insecurity issues in Ethiopia and Burkina Faso through the project Equip - Strengthening Smallholder Livestock Systems for the Future (grant number OPP11755487). These funds are administered by the Feed the Future Innovation Lab for Livestock Systems, which was established by funding from the United States Agency for International Development (USAID) and is co-led by the University of Florida’s Institute of Food and Agricultural Sciences and the International Livestock Research Institute. Support for the Feed the Future Innovation Lab for Livestock Systems is made possible by the generous support of the American people through USAID. The contents are the responsibility of the authors and do not necessarily reflect the views of USAID or the United States Government. REDCap is hosted at the University of Florida Clinical and Translational Science Institute (CTSI), supported by NIH National Center for Advancing Translational Sciences grant UL1 TR000064.

## Notes

### Competing Interest Statement

The authors have declared no competing interest.

### Author Declarations

Ethics approval and consent to participant. Ethical approval was obtained from the University of Florida Internal Review Board (IRB201903141); the Haramaya University Institutional Health Research Ethics Committee (COHMS/1010/3796/20) and the Ethiopia National Research Ethics Review Committee (SM/14.1/1059/20). Written informed consent is obtained from all participating households (husband and wife) using a form in the local language (Afaan Oromo). Research findings will be disseminated to community stakeholders, including participants, through the existing CAB. Findings will be disseminated to scientific, academic, policy and development stakeholders through conferences and peer-reviewed journals and through the Feed the Future Innovation Lab for Livestock Systems. The Bill and Melinda Gates Foundation, the funder of this trial, requires an open access data policy. Therefore, all manuscripts from this funded work will be open access with the data underlying the published research results available in a public repository. The website https://www.gatesfoundation.org/how-we-work/generalinformation/open-access-policy provides more information on this policy. A Community Advisory Board including a representative of the community, religious leaders (imam), woreda and kebele administration, woreda women and children affairs, woreda bureau of health and agriculture, kebele health, and agricultural extension workers was established to guide the research team for better understanding of local context and entry to the community and is regularly engaged in the research [20]. Only the project manager at Haramaya University and the data manager at the University of Florida have access to personally identifiable information in the REDCap database. Any data shared among researchers within the project was identified and blinded. Materials and Data Transfer Agreements assure confidentiality of data when exchanged with international partners and others.

